# First detection of Powassan Virus lineage I in field-collected *Dermacentor variabilis* from New York, USA

**DOI:** 10.1101/2022.03.01.22271704

**Authors:** Charles Hart, Erin Hassett, Chantal B.F. Vogels, Daniel Shapley, Nathan D. Grubaugh, Saravanan Thangamani

## Abstract

Powassan virus (POWV) is a tick-borne flavivirus that can cause lethal or debilitating neurological illness. It is canonically transmitted by *Ixodes* genus ticks but may interact with sympatric *Dermacentor* species. Here, we report the first detection of POWV lineage I from a pool of field-collected *D. variabilis* in New York state.

## INTRODUCTION

Powassan virus (POWV) is a neurotropic, tick-borne flavivirus first isolated in 1958 from the brain of a patient who had died of encephalitis (1). POWV infection results in febrile illness that may progress to encephalitis, meningitis, and, rarely, meningoencephalitis (2), which is associated with head pain, confusion, paralysis, coma, and death in up to 15% of cases. Additionally, more than half of survivors experience long-term neuro-sequelae including motor deficiency and cognitive deficits (3).

POWV was initially associated with the woodchuck tick, *Ixodes cookei* (4), although a secondary strain was also found in deer ticks (*Ixodes scapularis*) (5). This secondary virus was termed POWV lineage II, or deer tick virus (DTV). Due to the frequency with which *I. scapularis* bites humans compared to *I. cookei*, it is likely that DTV is the most common etiological source of Powassan encephalitis in the United States. However, this is difficult to discern due to serological homology between the virus lineages and the lack of viral genotyping in clinical settings.

Recently, interest has grown in the vector competency of other sympatric tick species. *Dermacentor spp*. ticks have been of particular interest due to their common occurrence in POWV/DTV-endemic areas and their tendency to bite humans. POWV has been isolated from *Dermacentor andersoni* in Colorado (6), with genetic analysis suggesting that this form, called POWV 791A-52, is most likely a form of DTV (7). However, it remains unclear if the tick in question was infected by spillover from another sylvatic cycle featuring *Ixodes* genus ticks or constitutes its own sylvatic system. Neither *scapularis* nor *I. cookei* are native to Colorado, although several rodent-specific *Ixodes* species are present (8) that may be involved in such a system.

Recent vector competency analysis has indicated that under laboratory conditions, *D. variabilis* is capable of acquiring and transmitting POWV, including maintaining replicating virus transstadially through molting (9). However, while this has been confirmed in experimentally infected ticks, it remains unclear if wild populations of *D. variabilis* can maintain and transmit POWV under natural circumstances. Considering that *D. variabilis* is the second most common human-biting species in New York (10), the ability for the species to transmit POWV in nature represents a critical component of potential human exposure.

Here, we report the first case of POWV detected and isolated from *D. variabilis* ticks.

## THE STUDY

As a part of our ongoing efforts to track the emergence of POWV in New York, we are performing tick surveillance in POWV hot-spots as identified from our community engaged tick testing program (10). In one of our hot-spots, we collected five adult female *D. variabilis* ticks, in addition to 68 *I. scapularis* ticks, in the latter half of April 2021, in Dutchess County, New York. These were visually speciated and assessed. The ticks were identified as unfed, female *D. variabilis*. These ticks were pooled, homogenized, and tested for the presence of POWV by rt-qPCR as described (10). Briefly, the presence of POWV was detected initially with a primer sensitive to both POWV lineage I and DTV. Following this, a differentiation qRT-PCR confirmed the detection of POWV lineage I with a titer of 3.88 Log10(FFU/ μgRNA). On the contrary, none of the *I. scapularis* collected from the same site tested positive for POWV lineage 1 and DTV.

We utilized our highly multiplexed PCR amplicon approach to sequence POWV detected from the tick homogenate (11). Libraries were prepared with the Illumina COVIDSeq Test RUO version, replacing the SARS-CoV-2 primers with POWV (12), and sequenced on the Illumina NovaSeq at the Yale Center for Genome Analysis. Consensus genomes were generated at a minimum nucleotide frequency threshold of 0.75 and minimum depth of 10 reads using iVar (version 1.3.1). We reconstructed a maximum-likelihood phylogenetic tree using IQ-TREE (version 1.6.12) with ultrafast bootstrap approximation (1000 replicates; 13). Our phylogenetic analysis revealed that the virus belongs to the POWV lineage I clade and is closely related to a POWV lineage I virus that we sequenced from *I. cookei* from New York in 2020 (**Fig. 1**). These two virus sequences represent the first time that POWV lineage I has been detected in the United States since 1964.

**Figure 1.**
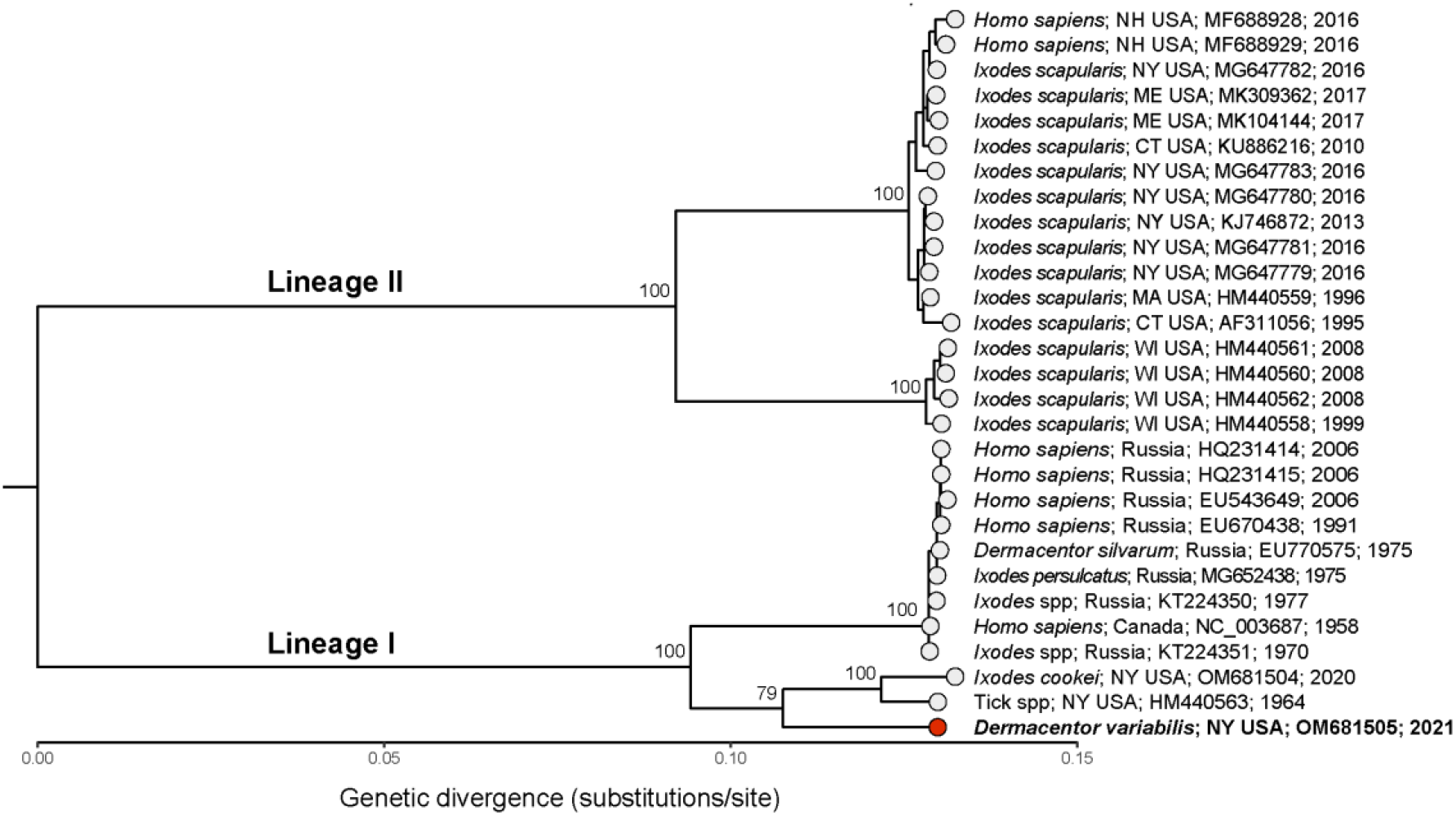
Maximum-likelihood phylogenetic tree of Powassan virus lineage I and II. We reconstructed phylogenetic relations using publicly available Powassan lineage I and II genomes. Highlighted in red is the virus isolated from *Dermacentor variabilis*, which is most closely related to other lineage I strains isolated from ticks in New York.

In addition, we used Sanger sequencing of tick ribosomal RNA to confirm that the sample was derived from *D. variabilis* and not another potential vector species. The resulting sequence had 100% homology to *D. variabilis* large subunit ribosomal RNA. Our results confirm the detection of POWV in *D. variabilis* in southern New York State, suggesting that POWV can exist in *D. variabilis*, either due to incidental exposure or due to its own sylvatic cycle. In this report, the POWV identified is similar to lineage I; this is normally associated with *I. cookei* and woodchucks (*Marmota monax*) instead of *I. scapularis* and *P. leucopus* (14). This suggests either spillover from that sylvatic cycle, a unique *D. variabilis-*dependent sylvatic cycle for POWV lineage I, or a unique subtype of the virus specific to *D. variabilis* with an unknown sylvatic cycle. Regardless of its source, our data indicate the possibility that some *D. variabilis* in POWV-endemic areas are capable of acquiring a strain of POWV similar to lineage I.

## CONCLUSIONS

Powassan virus is a medically important flavivirus understood to be primarily transmitted by *Ixodes* species in North America. It has recently been demonstrated, however, that the sympatric species *D. variabilis* is a competent vector for POWV under laboratory conditions (9). Here, we report the first detection of POWV lineage I in *D. variabilis* collected from the wild, suggesting that the species may play a direct role in POWV transmission in nature. Considering that *D. variabilis* is one of the primary human-biting ticks in New York, this presents a new potential source of human exposure to POWV.

## Data Availability

All data produced in the present work are contained in the manuscript.

## Author Bio

Dr. Charles Hart is a postdoctoral researcher at SUNY Upstate Medical University. His primary research focus is on the interaction between vector-borne pathogens, arthropod vectors, and vector-host interactions

